# Understanding quantity and intensity of hospital rehabilitation using electronic health record data

**DOI:** 10.1101/2023.05.25.23290461

**Authors:** Konstantin Georgiev, Dimitrios Doudesis, Joanne McPeake, Nicholas L Mills, Jacques Fleuriot, Susan D Shenkin, Atul Anand

**Affiliations:** BHF/University Centre for Cardiovascular Science, University of Edinburgh, UK; The Healthcare Improvement Studies Institute, University of Cambridge, UK; Artificial Intelligence and its Applications Institute, University of Edinburgh, UK; Ageing and Health, and Advanced Care Research Centre, Usher Institute, University of Edinburgh, UK

**Author notes:** **Corresponding author:** Dr Atul Anand, University of Edinburgh/BHF Centre for Cardiovascular Science, Room SU.226 Chancellor’s Building, 49 Little France Crescent, Edinburgh EH16 4SA, United Kingdom.

**Keywords:** hospital, rehabilitation, electronic health records

## Abstract

**Background:** Many hospitalised patients require rehabilitation during recovery from acute illness. We use routine data from electronic health records (EHR) to report the quantity and intensity of rehabilitation and compared this in patients with and without COVID-19.

**Methods:** We performed a retrospective cohort study of consecutive adults in whom COVID-19 testing was undertaken between March 2020 and August 2021 across three acute hospitals in Scotland. We defined rehabilitation contacts (physiotherapy, occupational therapy, dietetics and speech and language therapy) from timestamped EHR data and determined contact time from a linked workforce planning dataset. We included survivors to hospital discharge who received at least two rehabilitation contacts. The primary outcome was total rehabilitation time. Secondary outcomes included the number of contacts, time to first contact, and rehabilitation minutes per day. A multivariate regression analysis for predictors of rehabilitation time included age, sex, comorbidities, and socioeconomic status.

**Findings:** We included 11,591 consecutive unique patient admissions (76 [63,85] years, 56% female), of which 651 (6%) were with COVID-19, and 10,940 (94%) were admissions with negative testing. There were 128,646 rehabilitation contacts. Patients with COVID-19 received more than double the rehabilitation time compared to those without (365 [165,772] *vs* 170 [95,350] mins, p<0.001), and this was delivered over more specialist contacts (12 [6,25] *vs* 6 [3,11], p<0.001). Time to first rehabilitation contact was later in patients with COVID-19 (3 [1,5] *vs* 2 [1,4] days from admission). Overall, patients with COVID-19 received fewer minutes of rehabilitation per day of admission (14.1 [9.8,18.7] *vs* 15.6 [10.6,21.3], p<0.001). In our regression analyses, older age and COVID-19 were the most important predictors of increased rehabilitation time.

**Interpretation:** Patients with COVID received more rehabilitation contact time than those without COVID, but this was delivered less intensively. Rehabilitation data derived from the EHR represents a novel measure of delivered hospital care.

## Background

The delivery of hospital rehabilitation services during the COVID-19 pandemic presents a serious cause of concern. Survivors of infection have frequently required prolonged and complex rehabilitation. Throughout the pandemic, there has been a rapid change in the delivery of these services across hospitals and in the community. According to data synthesised by the Cochrane Rehabilitation Field,^1^ around 25-30% of people infected with COVID-19 in the early phases of the pandemic developed long-COVID with at least one persisting symptom at 6 months from the illness onset. These effects may be profound and impair more than just physical function.^2^ The impact of earlier targeting of rehabilitation on setting the trajectory of recovery remains unclear for those hospitalised with COVID-19, in part due to challenges in quantifying the amount of rehabilitation care delivered in this large patient population. The pathophysiology of these post-acute sequelae is also poorly understood, and their interactions in individuals are unpredictable.^3^

Rehabilitation guidelines are based on consensus with a focus on managing acute respiratory issues stemming from COVID-19.^4–6^ There is broad agreement that individualised treatment plans are required to adapt the delivery of rehabilitation according to factors such as patient frailty and premorbid function.^7^ Individualised rehabilitation plans are common outside of COVID-19, such as in hyperacute rehabilitation for patients with complex neurological disabilities or in the setting of major trauma.^8^ However, applying this approach to patients with COVID-19 would depend on reliable stratification to identify a population at risk of high rehabilitation needs. No previous studies have reported recovery trajectories in patients hospitalised with COVID-19 using systematic measures of rehabilitation delivery. However, many electronic health record (EHR) systems now capture routine data on rehabilitation, including timestamped logs of contacts with specialist providers. This presents new opportunities to understand the interventions delivered to patients and create a novel person-centred outcome measure to better appreciate the effects of acute illness – the specialist rehabilitation time required to achieve recovery to hospital discharge. Prior work using EHR data to understand rehabilitation has been limited to non-hospitalised populations.^9^ Other limitations include not capturing the full hospital admission,^10, 11^ or only involving selected types of therapy.^12^

In this study, we describe the rehabilitation needs of consecutive patients with and without COVID-19 infection across three acute hospitals, using routine EHR data to define the total quantity and intensity of care delivered.

## Methods

### Study design and participants

We conducted a retrospective cohort study of consecutive unique patients with an acute hospital admission between March 2020 and August 2021 to one of three hospitals in Scotland. Patients were included when at least one severe acute respiratory syndrome coronavirus-2 (SARS-CoV-2) test result was available using reverse transcriptase-polymerase chain reaction (RT-PCR). Admissions were classified as with COVID-19 in the event of any positive RT-PCR test during the admission period or in the ten days prior to admission from community testing. All other episodes with at least one negative RT-PCR result were designated as admissions without COVID-19. Individuals were only included for their first hospital admission with COVID-19 in the study period, and in the absence of any such admissions, only for their first hospital admission with negative testing. This created a study cohort of unique patients. Further inclusion criteria required at least two rehabilitation contacts within seven days of admission to identify a population truly engaged by rehabilitation teams rather than simply assessed once without ongoing needs. Patients were excluded from the study cohort if they died during their hospital stay to allow a clearer interpretation of rehabilitation received to achieve hospital discharge. Regardless of symptoms, widespread screening for SARS-CoV-2 was introduced for all hospitalised patients early in the pandemic, creating a large cohort of patients hospitalised without COVID-19.

### Linked data

The study was performed with the approval of the local Research Ethics Committee and delegated Caldicott Guardian in accordance with the Declaration of Helsinki. All data were collected from EHRs and national registries anonymised and linked by the DataLoch service (Edinburgh, United Kingdom) and analysed within their Secure Data Environment. Consent was not required or sought from individual patients. Baseline characteristics included age, sex, the Scottish Index of Multiple Deprivation (SIMD)^13^ quintile status and the presence of prior coded health conditions (including ischaemic heart disease, myocardial infarction, stroke, diabetes, obesity, dementia, delirium, depression, asthma and chronic obstructive pulmonary disease [COPD]). These morbidities were identified from national administrative records (Scottish Morbidity Record 01) covering prior hospitalisation with relevant ICD-10 codes, using code lists defined previously by the CALIBER research group for each listed condition.^14^ These features were also used as confounders for generating and testing an adjusted population using Inverse Probability Treatment Weighting (IPTW) (**appendix p 3, 9**).

Rehabilitation contact data were obtained from the EHR system (TrakCare, InterSystems, MA) in two formats: a timestamped log of any documented contact by a designated rehabilitation Allied Healthcare Professional (AHP), and a dedicated activity database quantifying the duration in minutes of treatment delivered to individual patients in each session. The activity database is used for workforce planning and service management. Four rehabilitation types were defined by the AHP providing input: physiotherapy, occupational therapy, speech and language therapy or dietetics. These databases were linked to produce a table of timestamped contacts by AHP type and minutes of therapy delivered for individual patients in the study cohort. Data imputation using the Moving Weighted Exponential Average (MWEA) was applied to handle any missingness within the recorded minutes of each intervention (**appendix p 4**).

### Outcomes

The primary outcome was total rehabilitation time, defined as the sum of all rehabilitation contact times across all AHP types, delivered between hospital admission and discharge. Secondary outcomes included the total number of rehabilitation contacts, time from admission to first or second consecutive rehabilitation contact, total rehabilitation time delivered out of hours (outside 7 AM to 7 PM), and mean rehabilitation time per day of hospital stay, as a proxy for rehabilitation intensity. Death from any cause at one year after hospital attendance was also determined by linkage with the National Records of Scotland.

### Statistical analysis

Due to asymptomatic screening, the large group of patients without COVID-19 (94% of admissions) warranted adjustment for potential confounders prior to hospitalisation. An IPTW-adjusted analysis was used to weigh the inverse probability of the exposure (COVID-19 status) given the patient characteristics at baseline.^15^ The modified population did not yield significant changes to the underlying model distribution, so this was not used in the primary analyses. However, IPTW-adjusted distribution plots were reported to attempt to adjust for any underlying confounders. Further analyses of the IPTW-generated cohorts are available in **appendix pp 6-8**.

Normally distributed variables are reported as mean ± SD, while non-normally distributed variables are described as median [IQR] and differences tested using parametric or non-parametric methods, respectively. For age-based analyses of rehabilitation time, we divided the population into adults below 65 years old (18-64 years) and then balanced the remaining older groups for size (65-82 years and >82 years). Density plots for total rehabilitation time were generated for these age groups and stratified by COVID-19 status. Length of stay was stratified by AHP intervention type to show differences between those receiving or not receiving at least one contact, regardless of COVID-19 status.

Additionally, we fitted a linear mixed effects regression (LMER) model, estimating predictors of total rehabilitation time (minutes), which was log-transformed due to skewness. We included the following covariates in this model: age, sex, SIMD, comorbidities prior to admission and COVID-19 status. Stepwise feature selection was applied after comparing selection methods and completing other preprocessing steps (described in **appendix pp 12-13**). The remaining covariates (with the resulting exclusion of stroke and COPD) were used to build a simple fixed-effects regression model, which was extended to include random effects terms to control for multi-level interactions that introduced large amounts of unexplained variance within the data. The final model adjusted for SIMD as the random intercept across age groups as the random slopes. The equations describing both fixed and mixed effect models are described in **appendix p 5**.

The benefits of incorporating random effects into the model fit were evaluated using the AIC score. Chi-squared testing indicated that a model incorporating random intercepts and slopes was more representative than one using only random intercepts (p<0.001). A restricted maximum likelihood (REML) optimisation function was used rather than the standard likelihood estimator due to potential bias in smaller sample sizes within the latter. To conform to the rules applied to random effect variables (particularly intercepts), we sought to include at least five levels, expanding the number of age groups and using SIMD estimates in quintiles. The overall fixed and mixed-effect performance metrics (marginal *R*^2^=0.363, conditional *R*^2^=0.822) represented the best fit to the observed data. Further testing of model performance and reliability is described in **appendix p 14**.

For the fixed effect variables, the unstandardised beta coefficients *b*_*i*_ were reported alongside their 95% confidence intervals (CI). All analyses were conducted in R (version 4.2.0), and significance was considered at p<0.05.

## Results

### Study population

There were 54,260 eligible index hospital episodes for patients with at least one valid COVID-19 test result between March 2020 and August 2021. This was restricted to an analysis cohort of 11,591 unique patients after excluding those who died during hospitalisation (n=4,852) or did not receive at least two rehabilitation contacts within seven days of admission (n=37,817). Further details of the data extraction and a summary of rehabilitation contacts data are shown in **Figure 1**. Within the analysis cohort, there were 651 (6%) and 10,940 (94%) patients with and without COVID-19, respectively.

**Figure 1.**
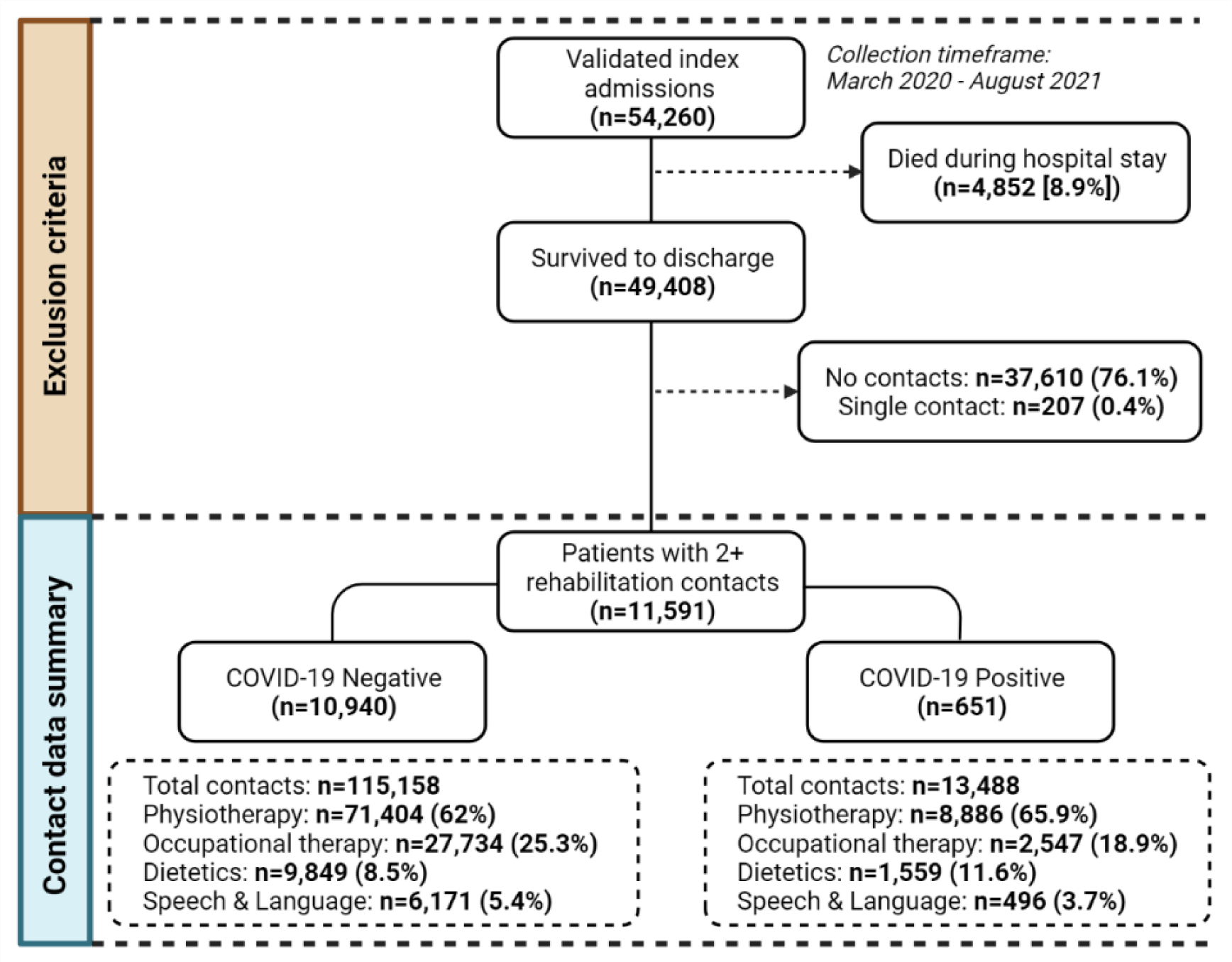
Summary of data flow into study.

Across these 11,591 hospital admissions, there were 128,646 rehabilitation contacts, of which 13,488 (11%) were delivered to patients with COVID-19. Physiotherapy accounted for nearly two-thirds of all rehabilitation interactions (n=80,290), with a higher proportion of patients with COVID-19 receiving this form of rehabilitation at least once compared to those without COVID-19 (66% *vs* 62%, p<0.001). The baseline characteristics of patients by COVID-19 status are shown in **Table 1**. Patients without COVID-19 were more likely to be female compared to those with COVID-19 (56% *vs* 50%, p=0.006), but there were no differences in age (76 [63, 85] *vs* 75 [60, 84], p=0.244) or history of common morbidities. The median length of hospital stay was more than doubled for patients with COVID-19 compared to those without (29 [14, 57] days *vs* 12 [7, 25] days, respectively, p<0·001). Death within one year of admission was lower in patients who had survived COVID-19 to hospital discharge, compared to those without infection (8% *vs* 12%, p=0.02).

**Table 1.** Baseline characteristics.

| Characteristic | All<br>(n=11,591) | COVID-19<br>negative<br>(n=10,940) | COVID-19<br>positive<br>(n=651) | p |
| --- | --- | --- | --- | --- |
| Age, years<br>(median, IQR) | 76 (63, 85) | 76 (63, 85) | 75 (60, 84) | 0.244 |
| Sex | .. | .. | .. | 0.006 |
| Male | 5,154 (44) | 4,831 (44) | 323 (50) | .. |
| Female | 6,437 (56) | 6,109 (56) | 328 (50) | .. |
| SIMD in quintiles | .. | .. | .. | 0.159 |
| 1 (most deprived) | 1,507 (13) | 1,416 (13) | 91 (14) | .. |
| 2 | 2,841 (25) | 2,666 (24) | 175 (27) | .. |
| 3 | 2,086 (18) | 1,986 (18) | 100 (15) | .. |
| 4 | 1,996 (17) | 1,875 (17) | 121 (19) | .. |
| 5 (least deprived) | 3,161 (27) | 2,997 (27) | 164 (25) | .. |
| IHD | 1,394 (12) | 1,324 (12) | 70 (11) | 0.304 |
| Stroke | 781 (7) | 744 (7) | 37 (6) | 0.269 |
| Myocardial infarction | 646 (6) | 618 (6) | 28 (4) | 0.145 |
| Diabetes | 1,322 (11) | 1,260 (12) | 62 (10) | 0.12 |
| Obesity | 422 (4) | 391 (4) | 31 (5) | 0.116 |
| Dementia | 459 (4) | 429 (4) | 30 (5) | 0.383 |
| Delirium | 963 (8) | 899 (8) | 64 (10) | 0.147 |
| Depression | 589 (5) | 561 (5) | 28 (4) | 0.351 |
| Asthma | 721 (6) | 679 (6) | 42 (7) | 0.801 |
| COPD | 1,028 (9) | 975 (9) | 53 (8) | 0.501 |
| Length of stay, days<br>(median, IQR) | 12.7 (7.0, 26.4) | 12.2 (6.9, 24.8) | 28.6 (13.9, 56.8) | <0.001 |
| Death at 1 year | 1,422 (12) | 1,367 (12) | 55 (8) | 0.02 |

Values are number (%) unless stated otherwise. SIMD – Scottish Index of Multiple Deprivation; IHD – Ischaemic Heart Disease; COPD – Chronic Obstructive Pulmonary Disease.

### Rehabilitation outcomes

The rehabilitation outcomes for the analysis population are shown in **Table 2**. The median total minutes of rehabilitation was more than doubled in patients with COVID-19 compared to those without (365 [165, 772] *vs* 170 [95, 350] minutes, p<0.001). Similarly, patients with COVID-19 received more rehabilitation contacts during their hospital admission (12 [6, 25] *vs* 6 [3, 11], p<0.001). Physiotherapy sessions were the most common intervention types in both groups (14±18 *vs* 6±10, p<0.001). However, as a result of the longer length of stay, the duration of rehabilitation per day was lower in patients with COVID-19 compared to those without (14.1 [9.8, 18.7] *vs* 15.6 [10.6, 21.3] mins, p<0.001). Patients with COVID-19 received their first and second rehabilitation contacts later than those without COVID-19 (3 [1, 5] *vs* 2 [1, 4] days from admission for the first contact; 5 [3, 7] *vs* 4 [2, 5] days for the second contact, respectively). There were no notable differences in the number of out-of-hours rehabilitation contacts.

**Table 2.** Rehabilitation outcomes.

| Characteristic | All<br>(n=11,591) | COVID-19<br>negative<br>(n=10,940) | COVID-19<br>positive<br>(n=651) | p |
| --- | --- | --- | --- | --- |
| Total minutes of rehabilitation | 175 (95, 370) | 170 (95, 350) | 365 (165, 772) | <0.001 |
| Number of interventions | 6 (3, 12) | 6 (3, 11) | 12 (6, 25) | <0.001 |
| Physiotherapy contacts (mean, SD) | 6.93 (10.82) | 6.53 (10.05) | 13.65 (18.43) | <0.001 |
| Occupational therapy contacts (mean, SD) | 2.61 (4.96) | 2.54 (4.9) | 3.91 (5.72) | <0.001 |
| Speech & Language therapy contacts (mean, SD) | 0.58 (3.16) | 0.56 (3.13) | 0.76 (3.57) | <0.001 |
| Dietetics contacts (mean, SD) | 0.98 (2.93) | 0.9 (2.78) | 2.39 (4.53) | <0.001 |
| Time to first contact (days) | 2 (1, 4) | 2 (1, 4) | 3 (1, 5) | <0.001 |
| Time to second 7-day contact (days) | 4 (2, 6) | 4 (2, 5) | 5 (3, 7) | <0.001 |
| Total minutes of out-of-hours rehabilitation (mean, SD) | 0.67 (6.99) | 0.66 (6.96) | 0.68 (7.54) | 0.642 |
| Minutes of rehabilitation per day of hospitalisation | 15.5 (10.45, 21.11) | 15.62 (10.55, 21.25) | 14.06 (9.83, 18.73) | <0.001 |

All values are median (IQR) unless specified otherwise. Out-of-hours treatment is defined as any rehabilitation contact between 7pm and 7am on any day.

Total rehabilitation time increased with age (**Figure 2**). This was most marked in those who also had COVID-19, such that the median total rehabilitation time in 18-64 year old patients with COVID-19 was longer than for those >82 years without COVID-19 (285 [130, 632] *vs* 205 [110, 405] mins, respectively). The number of pre-existing morbidities did not appear related to total rehabilitation time in those with or without COVID-19 (**appendix p 11**). The overall length of stay was longer in patients with COVID-19, and this variable was skewed with a long tail of prolonged admission times (**appendix p 10**). Receiving any type of AHP rehabilitation contact was associated with an extended hospital stay. This was most notable for patients that required speech and language therapy, where the median length of stay was 28 [12, 65] days compared to 12 [7, 23] days in those who did not receive this form of rehabilitation (p<0.001, **Figure 3**).

**Figure 2.**
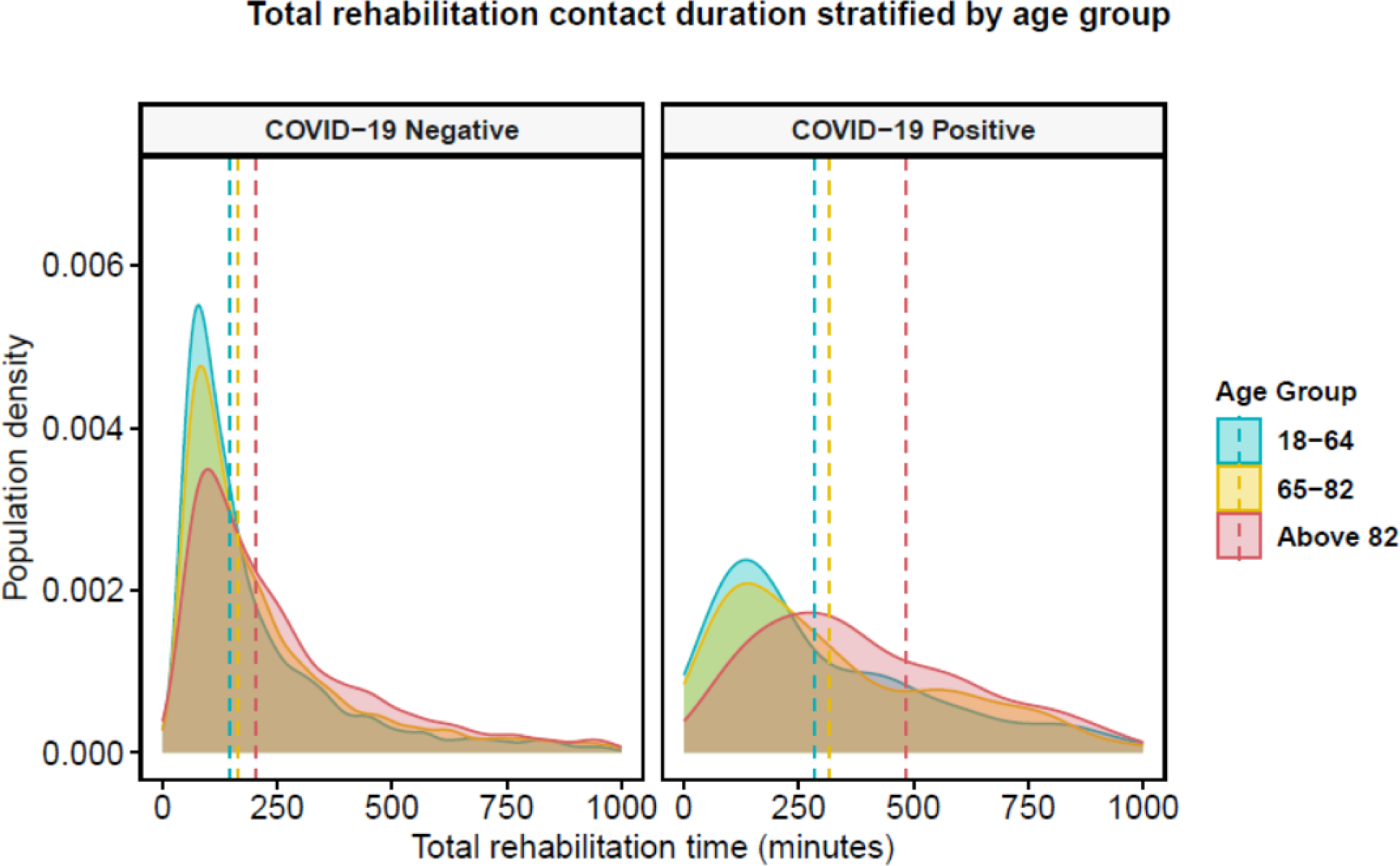
Distribution of IPTW-adjusted total rehabilitation time across the groups with and without COVID-19 stratified by three age categories. Vertical dashed lines represent the group median.

**Figure 3.**
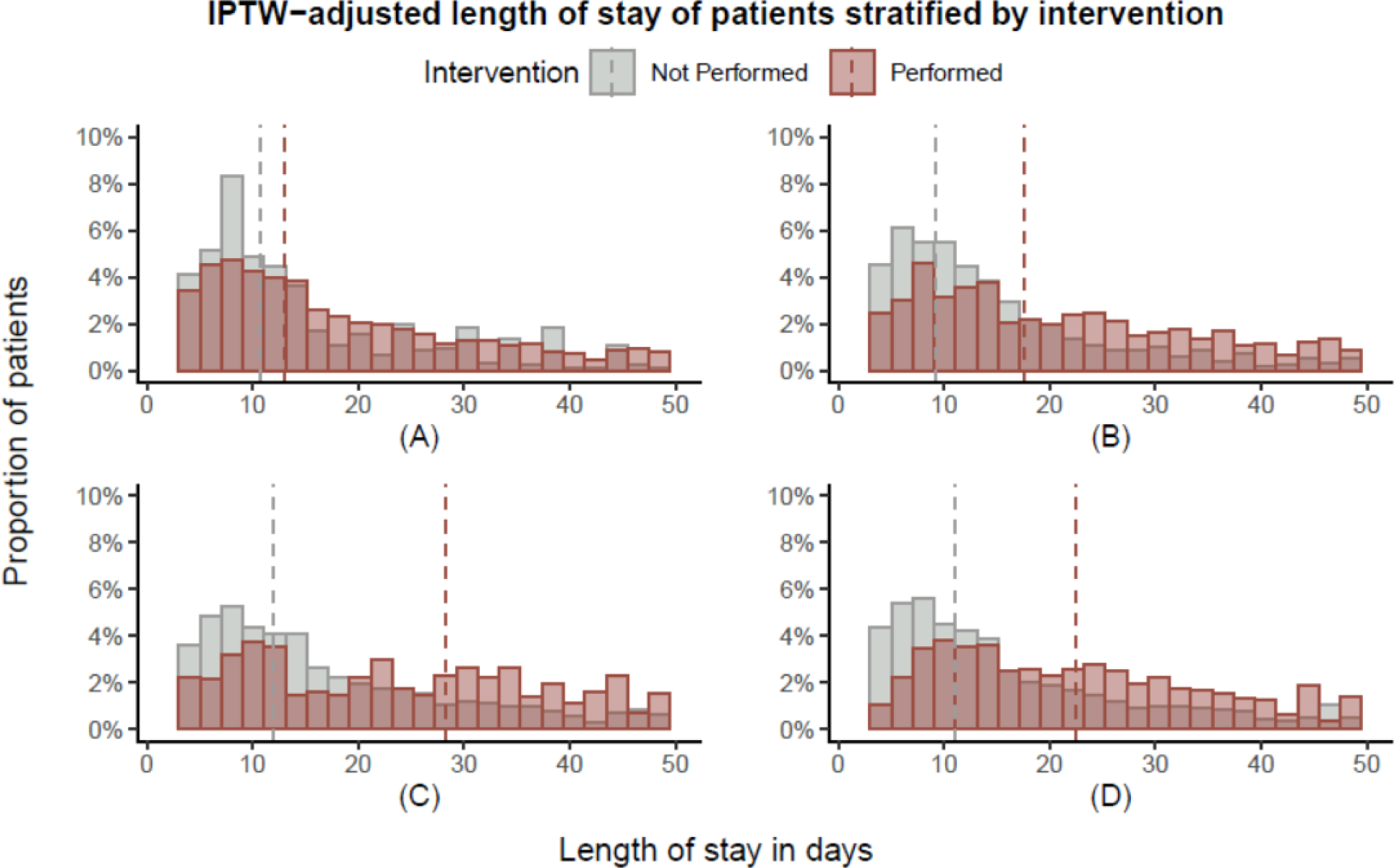
Distribution of the IPTW-adjusted hospital length of stay in days for patients who did and did not receive at least one specific AHP rehabilitation contacts: A – physiotherapy; B – occupational therapy; C – speech and language therapy; D – dietetics. Vertical dashed lines represent the group median.

### Regression analysis

The regression model outputs are summarised in **Table 3** (fixed effects) and **appendix p 14** (random effects). COVID-19 status was independently associated with increased total rehabilitation time (p<0.001), as was being in the oldest age group >79 years (p=0.03), but there were no differences by sex. The SIMD coefficients yielded mixed results using a reference group of the most deprived (quintile 1). There was no significant difference in association seen within the least deprived group (quintile 5), but individuals within quintiles 2 and 3 were more likely to receive higher rehabilitation time (p=0.001 and p=0.002, respectively). The correlation values between morbidities and rehabilitation time were small compared to the other covariates. Among the conditions included after stepwise selection, only a history of diabetes was associated with longer rehabilitation time (p=0.016). Other morbidities were negatively correlated with rehabilitation time, most notably for ischaemic heart disease (p<0.001), obesity (p<0.001) and asthma (p<0.001).

**Table 3.** Linear mixed effects regression model for predictors of total rehabilitation time

| log(Total minutes of rehabilitation) |  |  |  |  |
| --- | --- | --- | --- | --- |
| Predictors |  | Estimates | 95% CI | p |
| Age groups | 18-29 | Reference group | .. | .. |
|  | 30-49 | 0.08 | [-0.29, 0.45] | 0.662 |
|  | 50-65 | 0.17 | [-0.42, 0.76] | 0.563 |
|  | 66-79 | 0.2 | [-1.24, 1.65] | 0.782 |
|  | Above 79 | 0.6 | [0.06, 1.15] | 0.031 |
| Sex | Male | Reference group | .. | .. |
|  | Female | 0.02 | [-0.01, 0.05] | 0.144 |
| SIMD (Quintiles) | 1 (most deprived) | Reference group | .. | .. |
|  | 2 | 2.05 | [0.89, 3.2] | 0.001 |
|  | 3 | 2.09 | [0.8, 3.38] | 0.002 |
|  | 4 | -0.75 | [-2.01, 0.5] | 0.24 |
|  | 5 (least deprived) | -0.65 | [-1.87, 0.57] | 0.294 |
| Comorbidity | Ischaemic Heart Disease | -0.13 | [-0.18, -0.07] | <0.001 |
|  | Myocardial Infarction | -0.13 | [-0.22, -0.05] | 0.002 |
|  | Diabetes | 0.06 | [0.01, 0.1] | 0.016 |
|  | Obesity | -0.16 | [-0.24, -0.07] | <0.001 |
|  | Dementia | -0.09 | [-0.17, -0.02] | 0.015 |
|  | Delirium | -0.05 | [-0.1, 0.01] | 0.085 |
|  | Depression | -0.08 | [-0.15, -0.01] | 0.036 |
|  | Asthma | -0.17 | [-0.23, -0.11] | <0.001 |
| COVID-19 status | Negative | Reference group | .. | .. |
|  | Positive | 0.67 | [0.64, 0.69] | <0.001 |

Beta-coefficient estimates shown from a LMER model using SIMD as the random intercept and age group as the random slope terms. The reference groups for the comorbidities are patients without these conditions.

## Discussion

We report the rehabilitation needs of people hospitalised with and without COVID-19 using routine EHR data. Patients with COVID-19 received more than double the total rehabilitation contacts and time during their hospital stay compared to those with similar baseline characteristics without COVID-19. The total rehabilitation provision was greatest in the oldest patients with COVID-19. However, the rehabilitation time per day was lower for patients with COVID-19, implying a more prolonged recovery but with less intensive treatment. The first rehabilitation contact was also delivered later in those with COVID-19, with a possibility that infection control concerns or severe acute illness may have delayed the initiation of rehabilitation. Our work shows how these data can be successfully extracted from routine EHRs to provide a novel measure of patient recovery from acute hospitalised illness.

The association between severe infection with COVID-19 and increased rehabilitation needs is understood, although the quantity of these treatments and complex mechanisms driving prolonged recovery time in some individuals remains unclear.^3^ COVID-19 infection requiring hospital treatment is a clear risk factor for symptom persistence,^16, 17^ although much of this evidence comes from early waves of the pandemic in largely unvaccinated populations. Severe infection is a highly catabolic state.^18^ The loss of muscle mass in such patients would predictably increase the rehabilitation effort needed to regain sufficient physical function for hospital discharge. However, the effect of rehabilitation intensity early in the infective course is less clear. Our data suggest that those who survived COVID-19 received more rehabilitation overall, but this was spread over more prolonged hospital admissions and was initiated later in the hospital stay. As an observational study, we can only speculate on the direction of the relationship between rehabilitation access and length of hospital stay. It is plausible that COVID-19 patients had limited potential to engage in physical therapy, either because of illness acuity or post-infection complications.

Early access to rehabilitation following acute illness has been associated with lower mortality rates and better functional outcomes in other non-COVID illnesses.^19, 20^ Our data showed a comparative delay of one day in the receipt of a first rehabilitation contact for patients with COVID-19, compared to similar individuals without infection. It is possible that isolation and barrier nursing requirements of infected patients may have limited early access to therapy, which might partly explain the less intensive rehabilitation delivered. There is some evidence for focused or ‘hyperacute’ rehabilitation in dedicated units for COVID-19, with one small study of 100 patients reporting improved recovery in respiratory and muscle function.^21^ However, scaling such services to a vast post-COVID population is challenging. For future infection waves or pandemics, routine EHR data could help identify patients with high predicted rehabilitation needs and focus limited resources on a smaller target population more likely to benefit from intensive treatment.

Even before the pandemic, ‘hospital-acquired disability’ was growing in attention, with evidence that over a third of older hospitalised patients developed worsening disability by the time of discharge.^22, 23^ Our attempts to model the predictors of increased rehabilitation need were limited by the patient variables available in our dataset, but suggested that the oldest patients with higher socioeconomic deprivation required more total rehabilitation time. Those with existing comorbidities tended to receive less rehabilitation in our study. This may initially appear counterintuitive but could reflect a lower target functional level for hospital discharge in these patients. It is also possible that a reduction in function to a new lower baseline is accepted more readily in those with greater medical comorbidity, while more independent patients with better premorbid function may have been given a greater chance to improve with prolonged rehabilitation. Therapeutic nihilism is well recognised in the rehabilitation of patients with dementia,^24^ despite increasing evidence that meaningful gains can be achieved using tailored techniques.^25^

To our knowledge, this is the first study reporting routine AHP specialist contact data from consecutive patients to determine delivered rehabilitation time using the extensive timestamped data available within modern EHR systems. Length of stay is regularly reported as a measure of care quality, with an assumption that effective health services discharge patients from acute services earlier. However, it is also well-recognised that a patient’s length of stay is determined by multiple non-clinical factors, including the availability of social care services or access to post-acute care units.^26^ More fundamentally, using the length of stay as a proxy outcome measure for successful rehabilitation fails to capture the actual treatment delivered to patients. In the United Kingdom, NHS Improvement has developed a visual management system to show ‘red’ and ‘green’ days during a patient’s hospital admission. Red days are where a patient receives little or no value-added care, whereas green days reflect some input to progress a patient towards discharge.^27^ In one study reviewing a single acute geriatric unit, 54% of admission days were classified as ‘red’, with most of the inactivity explained by delays waiting for community services.^28^ Whilst this approach may provide valuable data to focus attention on system weaknesses in delivering efficient acute care, it currently requires manual review of every occupied bed in a hospital unit using a spot audit technique. This is highly inefficient and difficult to scale to a whole hospital or healthcare system. EHR measures of healthcare professional contacts represent a novel approach to reviewing system inactivity in near real-time.

There are several strengths to our study. We were able to include a large cohort of consecutive patients across multiple acute hospitals due to a shared EHR system. By limiting our analysis to patients with multiple rehabilitation contacts, we created groups of patients with and without COVID-19 who had similar baseline characteristics. By using an integrated health provider dataset, we were able to include contacts from across the entire admission period, including step-down rehabilitation units that are often excluded from hospital-based studies focused only on acute care. However, as a retrospective cohort study, we must acknowledge some limitations to our approach. We can only speculate on the content or the effect of rehabilitation received. Our data show the rehabilitation received by patients but cannot determine unmet needs, which might have varied with clinical pressures during the pandemic. We required some imputation for our rehabilitation time variable, reflecting our use of a workforce planning database which included missing data. The COVID-19 status of our cohort was determined by laboratory testing results and not by clinical status. It is likely that some patients in our group with a positive COVID-19 test were minimally affected by the infection, particularly in the later period of analysis where acquired population immunity had increased. For this analysis, we had access to limited comorbidity information from previous hospitalisation data, and this would be strengthened by wider data linkage, particularly for primary care records.

In conclusion, we have extracted rehabilitation information from hospital EHRs, showing the increased needs of patients with COVID-19 for prolonged care. Our approach offers an alternative method to understand the complexity of recovery after acute illness, which could form a novel evaluation endpoint for future intervention studies. Rehabilitation contacts and time provides more context to acute hospital care than widely reported measures such as length of hospital stay.

### Availability of data and materials

This study makes use of several routine electronic health care data sources that are linked, de-identified, and held in a Secure Data Environment (DataLoch), which is accessible by approved individuals who have undertaken the necessary governance training. Summary data can be made available upon request to the corresponding author. All data processing and modelling were conducted in RStudio version 4.2.0 using publicly available libraries through CRAN. The data extraction and cleaning procedures were performed using *tidyverse*, the visualisations were performed using *ggplot2*, the mixed-effects model was fit using *lme4* and the weights for the IPTW adjustments were generated using the *ipw* and *survey* packages. The data analysis code is held in a Secure Data Environment but may be made available for review in a non-disclosive format on discussion with the corresponding author.

## Data Availability

This study makes use of several routine electronic health care data sources that are linked, de-identified, and held in a Secure Data Environment (DataLoch), which is accessible by approved individuals who have undertaken the necessary governance training. Summary data can be made available upon request to the corresponding author. All data processing and modelling were conducted in RStudio version 4.2.0 using publicly available libraries through CRAN. The data extraction and cleaning procedures were performed using tidyverse, the visualisations were performed using ggplot2, the mixed-effects model was fit using lme4 and the weights for the IPTW adjustments were generated using the ipw and survey packages. The data analysis code is held in a Secure Data Environment but may be made available for review in a non-disclosive format on discussion with the corresponding author.

## Author contributions

KG and AA conceived the study and its design. KG performed the analysis. DD assisted in refining the methodology. KG and AA had access to and verified the raw data and analysis tables. KG, AA and DD interpreted the data. KG drafted the manuscript. AA, NM, JM, SS and JF revised the manuscript critically for important intellectual content. All authors provided their final approval of the version to be published. All authors are accountable for the work.

## Ethics declarations

### Ethics approval and consent to participate

This work was reviewed by the DataLoch ethics review panel (LPAC) and approved under Lothian REC approval to this service (Edinburgh, United Kingdom, ref: 17/NS/0072). All data was collected from Electronic Health Records and national registries previously anonymised by the DataLoch service and analysed in a Secure Data Environment. Consent from patients was not required for this study.

### Consent for publication

Not applicable

