## Supplement Document for "Understanding quantity and intensity of hospital rehabilitation using electronic health record data"

**Short Title:** Understanding hospital rehabilitation using electronic health records

**Corresponding author:**

Dr Atul Anand

University of Edinburgh/BHF Centre for Cardiovascular Science

Room SU.226 Chancellor’s Building

49 Little France Crescent

Edinburgh EH16 4SA

United Kingdom

### I. **Supplementary Methods**

#### 1. **Generating an IPTW-adjusted weighted population relative to COVID-19 status**

The application of Inverse Probability of Treatment Weighting (IPTW) produced a balanced sample size between the two groups by setting larger weights to exposed individuals with a low likelihood of being COVID-19 positive (and unexposed patients with a high probability of COVID-19). Within the weighted population, the patient characteristics were standardised by bootstrapping the samples for these specific minority groups.^1^ Rather than using 1:1 matching (as in Propensity Score Matching [PSM]) and eliminating a significant subset of the control samples,^2,3^ this approach enabled us to use appropriately weighted proportions of the entire set of patients. IPTW algorithms estimate the Average Treatment Effect (ATE) across the full study population, focusing on the complex samples of the individual groups (COVID-19 positive and COVID-19 negative), which are weighted corresponding to the population-level.^4^

The metric used to estimate the group balance was the feature-wise Standardised Mean Difference score (SMD) shown for the data before and after adjustment. SMD values below 0.1 were deemed satisfactory for the adjustment algorithm, consistent with other literature.^3,5^ However, due to the subjective nature of adjusting for covariates from different data sources, we sought to choose the balancing algorithm that minimised this score as much as possible. The Wilcoxon rank-sum test for complex survey weights was used to estimate the p-values in continuous data.^6^ The chi-squared test with second-order correction was applied for categorical adjusted data.^7^

#### 2. **Data imputation of the missing specialist contact durations**

Various data imputation techniques were applied to produce an unbiased estimate of the missing AHP contact durations extracted from the routine data. The total degree of missingness was 33·1% of all relevant contact samples. Several time-series imputation techniques (along with simple imputations such as mean and median population mean) were compared. There seemed to be an association between the degree of missingness and the recording date. The results can be seen in Figure S1. The recorded minutes in each session were all a factor of five in the original data. Hence, a rounded-down moving average (RDOWN:MWEA) estimate proved to be the most accurate, achieving a Mean Absolute Error (MAE) of 17·09 and a Root Mean Squared Error (RMSE) of 24·51. As a result, this technique was chosen for imputation, which closely resembled the original distribution of contacts.

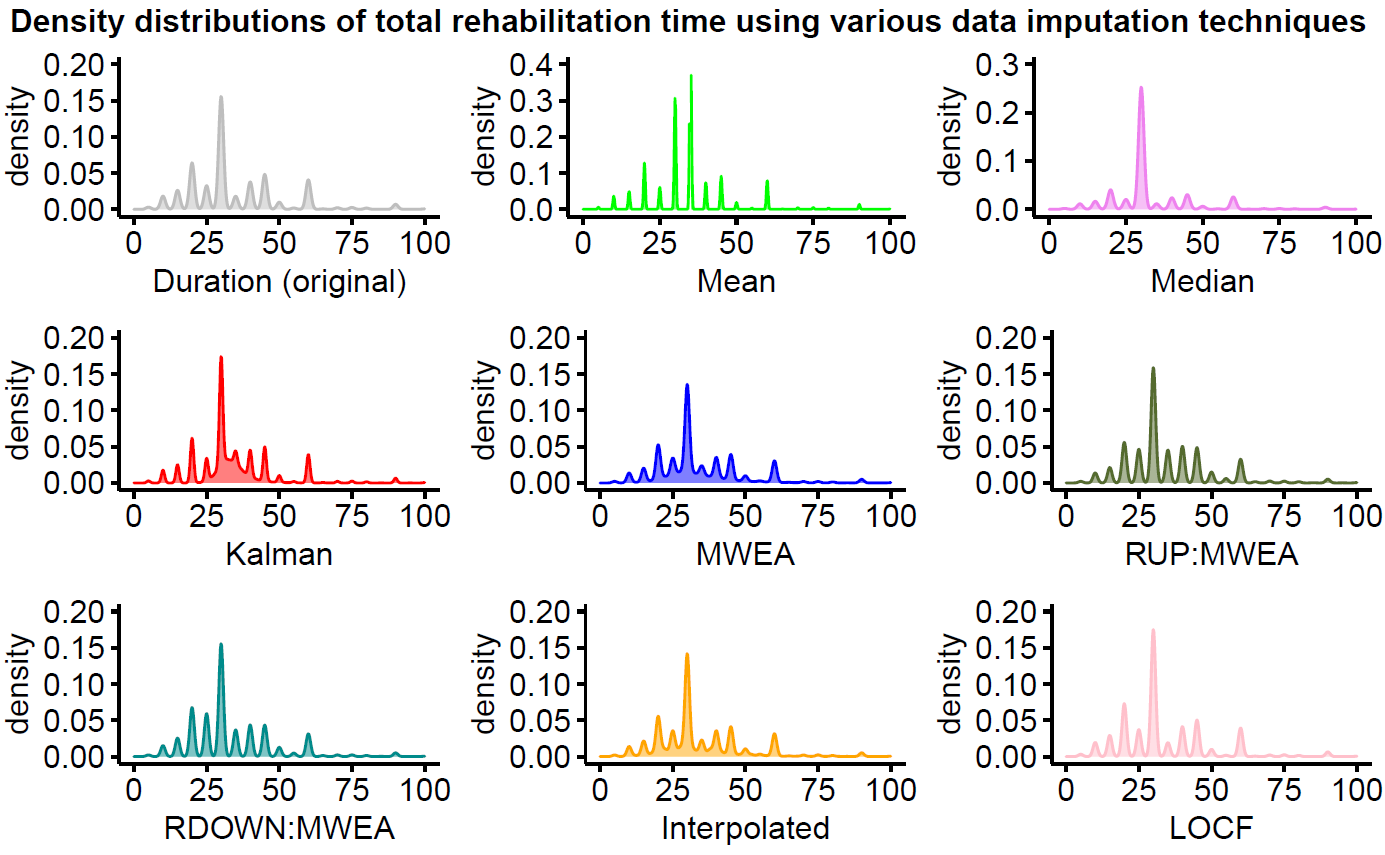

**Figure S1. Data imputation techniques used to approximate contact session duration (defined from left to right, top to bottom): gray – original distribution (no imputation); light green – all samples imputed with population mean; pink – all samples imputed with population median, red – time-series based Kalman smoothing; blue – moving weighted exponential average; olive green – rounded up moving weighted exponential average (by factor of 5); dark cyan – rounded down moving exponential average (by factor of 5); orange – time-series based linear interpolation; violet - last observation carried forward;**

#### 3. **Fixed and mixed regression representations**

The initial fixed-effects regression model (Ordinary Least Squares model [OLSR]) was expressed as:

$$\log\left( f_{x} \right)=\alpha+\beta_{1}x_{i}+ \beta_{2}x_{i+1}+\ldots+ \beta_{n}x_{i+n}+\epsilon$$

- where, α represents the intercept term
- $\beta_{i}x_{i}$ are the respective slopes and values across each covariate chosen using stepwise selection (included variables are age group, sex, SIMD, coronary heart disease, myocardial infarction, diabetes, obesity, dementia, delirium, depression and asthma)
- $\epsilon$ is the error term
- $log(f_{x})$ is the log-transformed total minutes of rehabilitation

The optimised mixed-effects model (LMER) was represented as follows:

$$\log\left( f_{x} \right)=\alpha+\beta_{1}x_{i}+ \beta_{2}x_{i+1}+\ldots+ \beta_{n}x_{i+n}+b_{r0}+b_{r1}x_{r1}+\epsilon$$

- where, $b_{r0}$ is the random intercept term, the SIMD in quintiles
- $b_{r1}x_{r1}$ represents the random slope term $b_{r1}$ for age group, modifying the subject $x_{r1}$

Model variations for both were fit on the transformed version of the dataset, described in **Table S3**. The fixed effects 95% Confidence Intervals (CIs) and p-values of the adapted variables were computed using a Wald t-distribution approximation. The resulting model was evaluated for signs of heteroscedasticity (an increase of variance in the residuals alongside the rise of values in the response variable) using the Breusch-Pagan test.^8^ Signs of model singularity were also assessed, where the variance-covariance matrix of the model would become zero-inflated due to high model complexity and insufficient samples.^9^

### II. **Supplementary results**

#### 1. **IPTW-adjusted baseline characteristics**

The baseline characteristics were observed including the IPTW-adjusted population (Table S1). Only the SMD scores between groups for sex (0·11) and SIMD (0·1) were above the pre-determined difference threshold from the literature. However, the list of confounders included all listed variables except for length of stay and mortality. After adjustment, all characteristics had differences significantly closer to zero. Significant differences in sex (p=0·006) regarding the larger prevalence of COVID-negative female patients were adjusted for. After adjustment, the difference in all-cause mortality at one year was more significant (p=0·001). However, the differences length-of-stay remained similar.

**Table S1. Baseline characteristics with included IPTW adjustment results. Values are number (%) unless stated otherwise. Percentages are not shown for the IPTW-adjusted sample as they do not reflect the true distribution in a pseudo-cohort. SIMD – Scottish Index of Multiple Deprivation; IHD – Ischaemic Heart Disease; COPD – Chronic Obstructive Pulmonary Disease.**

| Characteristic | Unadjusted (original cohort) | | | | IPTW-adjusted (pseudo-cohort) | | | |
| --- | --- | --- | --- | --- | --- | --- | --- | --- |
|  | **COVID negative (n=10,940)** | **COVID positive (n=651)** | **p** | **SMD** | **COVID negative (n=11,591)** | **COVID positive (n=11,608)** | **p** | **SMD** |
| Age, years  (median, IQR) | 76 (63, 85) | 75 (60, 84) | 0.244 | 0.02 | 76 (63, 85) | 76 (61, 85) | 0.51 | 0.01 |
| Sex | .. | .. | 0.006 | 0.11 | .. | .. | 0.88 | 0.01 |
| Male | 4,831 (44) | 323 (50) | .. | .. | 6,437 | 6,481 | .. | .. |
| Female | 6,109 (56) | 328 (50) | .. | .. | 5,154 | 5,125 | .. | .. |
| SIMD in quintiles | .. | .. | 0.159 | 0.1 | .. | .. | >0.999 | <0.001 |
| 1 (most deprived) | 1,416 (13) | 91 (14) | .. | .. | 1,507 | 1,544 | .. | .. |
| 2 | 2,666 (24) | 175 (27) | .. | .. | 2,841 | 2,815 | .. | .. |
| 3 | 1,986 (18) | 100 (15) | .. | .. | 2,086 | 2,074 | .. | .. |
| 4 | 1,875 (17) | 121 (19) | .. | .. | 1,996 | 1,984 | .. | .. |
| 5 (least deprived) | 2,997 (27) | 164 (25) | .. | .. | 3,161 | 3,189 | .. | .. |
| IHD | 1,324 (12) | 70 (11) | 0.304 | 0.04 | 1,394 | 1,407 | 0.945 | <0.001 |
| Stroke | 744 (7) | 37 (6) | 0.269 | 0.05 | 781 | 786 | 0.974 | <0.001 |
| Myocardial infarction | 618 (6) | 28 (4) | 0.145 | 0.06 | 646 | 673 | 0.837 | 0.01 |
| Diabetes | 1,260 (12) | 62 (10) | 0.12 | 0.07 | 1,322 | 1,398 | 0.662 | 0.02 |
| Obesity | 391 (4) | 31 (5) | 0.116 | 0.06 | 422 | 437 | 0.857 | 0.01 |
| Dementia | 429 (4) | 30 (5) | 0.383 | 0.03 | 459 | 479 | 0.825 | 0.01 |
| Delirium | 899 (8) | 64 (10) | 0.147 | 0.06 | 963 | 950 | 0.907 | <0.001 |
| Depression | 561 (5) | 28 (4) | 0.351 | 0.04 | 589 | 586 | 0.97 | <0.001 |
| Asthma | 679 (6) | 42 (7) | 0.801 | 0.01 | 721 | 749 | 0.817 | 0.01 |
| COPD | 975 (9) | 53 (8) | 0.501 | 0.03 | 1,028 | 1,054 | 0.865 | 0.01 |
| Length of stay, days (median, IQR) | 12.2 (6.9, 24.8) | 28.6 (13.9, 56.8) | <0.001 | -0.51 | 12.2 (6.9, 24.8) | 28.6 (14.0, 56.8) | <0.001 | -0.52 |
| Death at 1 year | 1,367 (12) | 55 (8) | 0.02 | 0.13 | 1,450 | 964 | 0.001 | 0.14 |

#### 2. **IPTW-adjusted rehabilitation outcomes**

The differences in rehabilitation outcomes were also observed after adjustment. The IPTW algorithm did not have a visible impact on any of the rehabilitation requirements (Table S2). All relevant variables, relative to minutes of rehabilitation, number of interventions, time to contact and amount of rehabilitation per day, were mostly unchanged and remained significant after adjustment (p<0·001).

**Table S2. Rehabilitation outcomes with included IPTW adjustment results. All values are median (IQR) unless stated otherwise. Out-of-hours treatment is defined as any rehabilitation contact between 7pm-7am on any day.**

|  | Unadjusted (original cohort) | | | | IPTW-adjusted (pseudo-cohort) | | | |
| --- | --- | --- | --- | --- | --- | --- | --- | --- |
| Characteristic | **COVID-19 negative (n=10,940)** | **COVID-19 positive (n=651)** | **p** | **SMD** | **COVID-19 negative (n=11,591)** | **COVID-19 positive (n=11,608)** | **p** | **SMD** |
| Total minutes of rehabilitation | 170 (95, 350) | 365 (165, 772) | <0.001 | -0.43 | 170 (95, 350) | 362 (165, 775) | <0.001 | -0.43 |
| Number of interventions | 6 (3, 11) | 12 (6, 25) | <0.001 | -0.5 | 6 (3, 11) | 12 (6, 25) | <0.001 | -0.5 |
| Physiotherapy contacts (Mean, SD) | 6.53 (10.05) | 13.65 (18.43) | <0.001 | -0.48 | 6.53 (10.06) | 13.61 (18.22) | <0.001 | -0.48 |
| Occupational therapy contacts (Mean, SD) | 2.54 (4.9) | 3.91 (5.72) | <0.001 | -0.26 | 2.53 (4.91) | 3.99 (5.76) | <0.001 | -0.27 |
| Speech & Language therapy contacts (Mean, SD) | 0.56 (3.13) | 0.76 (3.57) | <0.001 | -0.06 | 0.56 (3.13) | 0.74 (3.44) | <0.001 | -0.05 |
| Dietetics contacts (Mean, SD) | 0.9 (2.78) | 2.39 (4.53) | <0.001 | -0.4 | 0.9 (2.78) | 2.37 (4.47) | <0.001 | -0.39 |
| Time to first contact (days) | 2 (1, 4) | 3 (1, 5) | <0.001 | -0.16 | 2 (1, 4) | 3 (1, 5) | <0.001 | -0.17 |
| Time to second 7-day contact (days) | 4 (2, 5) | 5 (3, 7) | <0.001 | -0.19 | 4 (2, 5) | 5 (3, 7) | <0.001 | -0.19 |
| Total minutes of out-of-hours rehabilitation (Mean, SD) | 0.66 (6.96) | 0.68 (7.54) | 0.642 | <0.001 | 0.66 (6.96) | 0.74 (8.01) | 0.552 | -0.01 |
| Minutes of rehabilitation per day of hospitalisation | 15.62 (10.55, 21.25) | 14.06 (9.83, 18.73) | <0.001 | 0.29 | 15.62 (10.53, 21.25) | 14.04 (9.94, 18.67) | <0.001 | 0.3 |

#### 3. **Performance analysis of cohort matching algorithms**

A sensitivity analysis was performed on common propensity-score matching (PSM) and IPTW algorithms with different variations. Statistical sampling techniques were evaluated to minimise the SMD score of the confounders defined at baseline (Figure S2). In the unmatched distribution, there was a significant difference between the covariates describing sex and SIMD between the treated and control groups. PSM was tested with three 1:1 matching techniques, which discarded a significant proportion of the control samples (COVID-19 negative patients) to generate a balanced population. The IPTW techniques utilised survey weights to resample both populations based on their correlation score with the exposure to COVID-19, generating a pseudo-cohort of weighted cases. This method was preferred as it did not discard any samples from the original cohort. IPTW with truncation at the 99^th^ percentile was also tested. However, this added more noise to the input weights. The best performance achieved across all covariates was reached by the genetic distance PSM, closely followed by the IPTW standard algorithm. The latter was chosen for the analysis based on its potential benefits in including a wider global population.

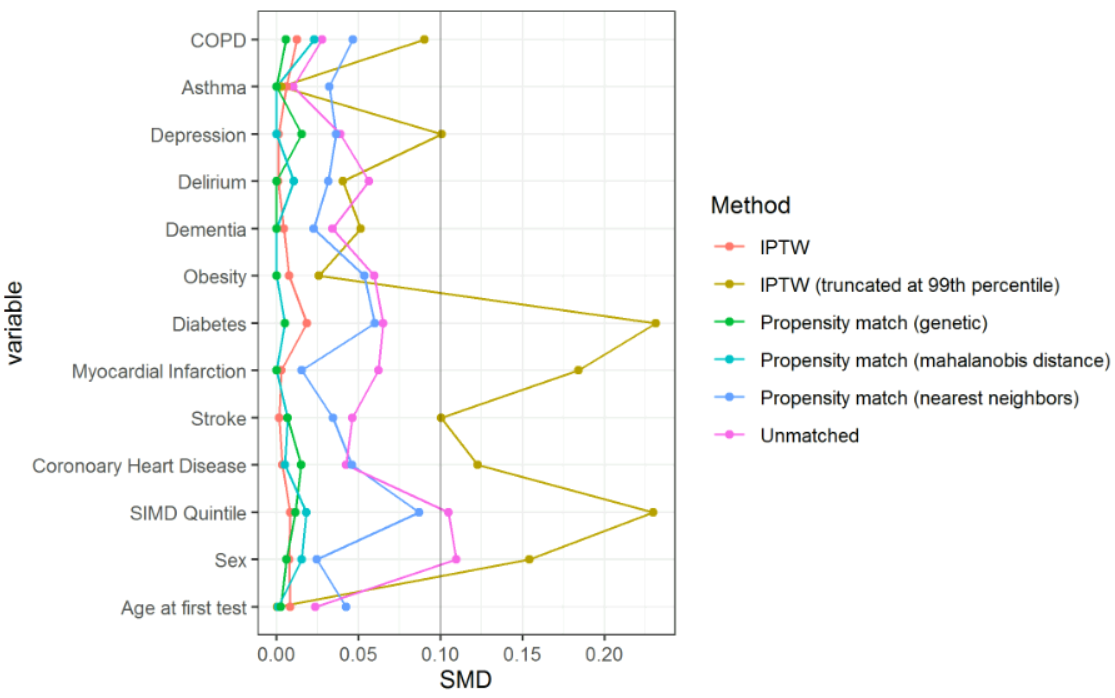

**Figure S2. Balancing effects of matching subsamples using the PSM and IPTW techniques on the target COVID-19 population; Gray vertical line represents the cutoff threshold of 0.1 expressed in the literature**

#### 4. **Adjusted patient length of stay density plot**

The observed COVID-19-positive patients were characterised by an extended length of stay, as seen in Figure S3**.** After IPTW adjustment, COVID-19-positive patients with a second contact had a considerably higher median length of stay (WMD=22·5 vs 11·57). The maximum length of stay of the compared distributions reached 76 days among the COVID-19-positive group compared to 42 days among their COVID-19-negative counterparts.

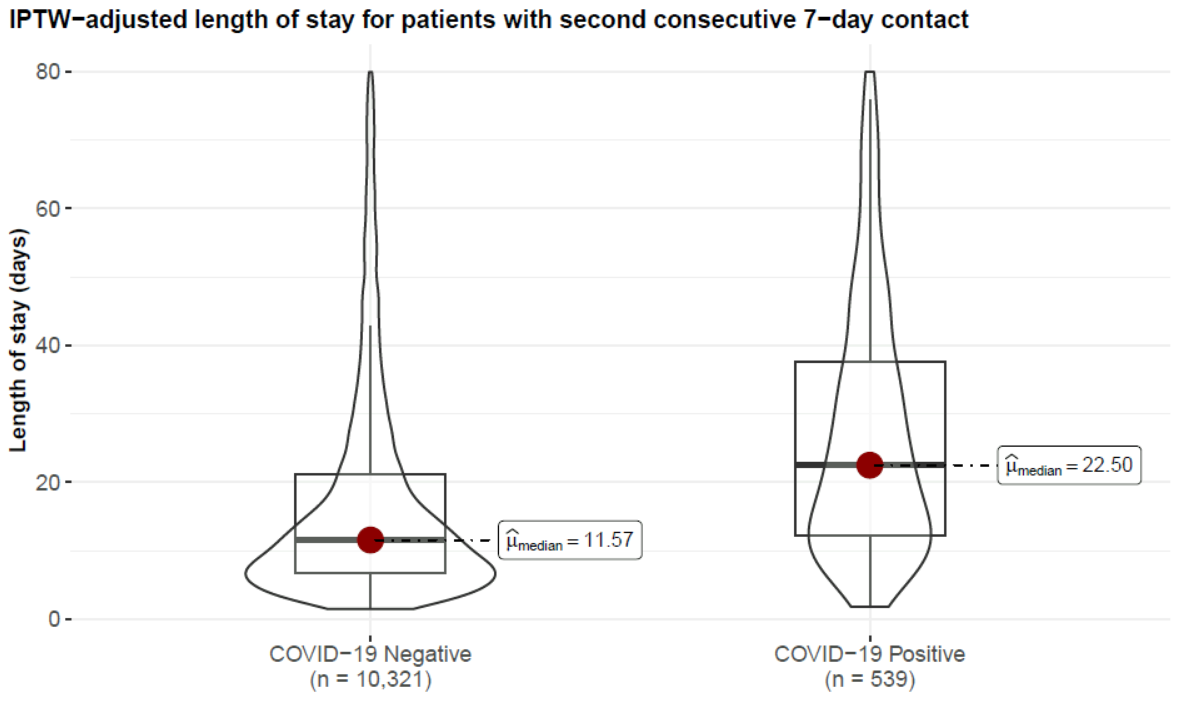

**Figure S3. Bar and violin plot highlighting the IPTW-adjusted length-of-stay between COVID-19 positive and negative patients with recorded second 7-day therapy contact; Samples are trimmed for outliers retaining the 90^th^ percentile of the distribution**

#### 5. **Rehabilitation time by number of premorbid conditions**

Figure S4 presents a patient density plot stratified by the number of comorbidities (before admission). The conditions used to estimate this number were based on the list of collected CALIBER phenotype codes. The results were compared between COVID-19-positive and COVID-19-negative hospitalised patients. As indicated by the areas, nonmorbid, morbid and multimorbid patients were not significantly different in terms of rehabilitation time. The median values were also similar in terms of morbidity. However, COVID-19-treated patients had a significantly higher value. The small sample size of the CALIBER codes and the small lookup timeframe (one year) is a limitation of these observations.

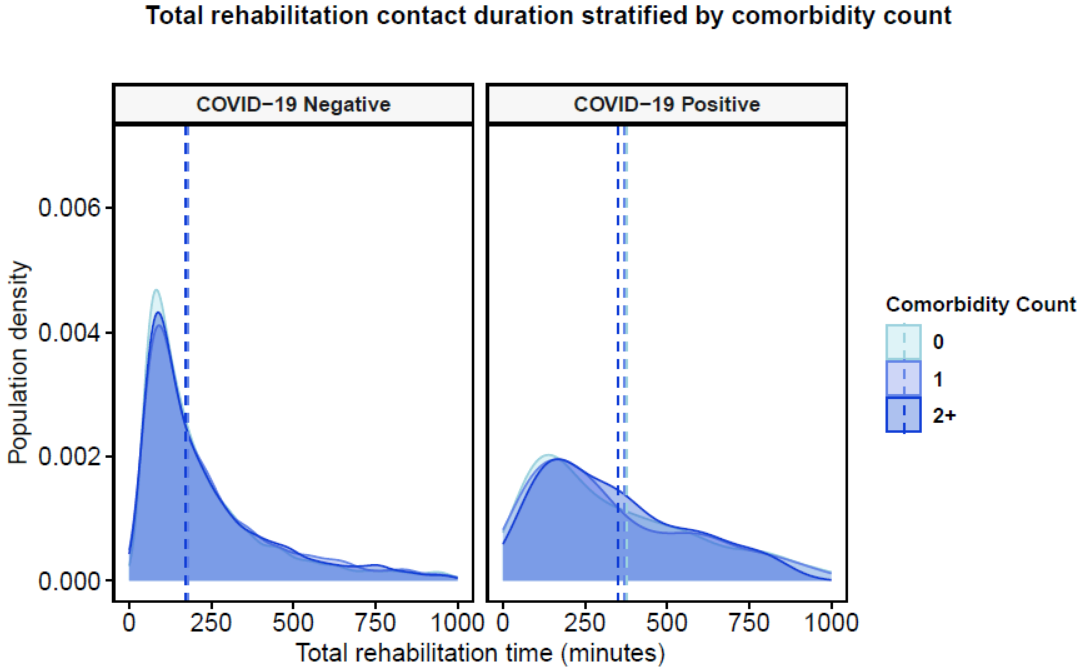

**Figure S4. Total rehabilitation time stratified by exposure and number of recorded premorbid conditions via CALIBER phenotyping; Vertical dashed lines highlight the group median;**

#### 6. **Optimising the dataset and tuning the random effect terms**

The results from the model sensitivity analysis, highlighting the chosen mixed effects model, are provided in Table S3. The OLSR was initially fitted using the covariates for age, sex, SIMD, COVID-19 status and the complete list of comorbidities estimating the log-transformed outcome of total rehabilitation time in minutes. The model was evaluated on the original set of 11,591 patients (10,940 COVID-19-negative and 651 COVID-19-positive) and yielded poor results relative to the explained variance. Afterwards, COVID-19-positive samples were repeatedly oversampled at random to balance the dataset for COVID-19 status, generating a total of 21,798 samples and improving the overall measure of fit. After removing outliers with a significant influence measure, according to Cook’s distance,^10^ the $R^{2}$ score was improved, and the error and quality measures (Akaike Information Criterion [AIC] and Bayesian Information Criterion [BIC]) were significantly reduced. Finally, a stepwise feature selection method was used to drop any significantly correlated input covariates by a method of sequential replacement relative to the AIC score.^11^ Forward and backward elimination were also tested, yielding similar results for this distribution. The stepwise selection algorithm removed the presence of stroke and COPD from the list of covariates, retaining the overall performance.

The resulting OLSR model was modified by introducing random intercept and slope interaction terms. As random intercepts should generally represent multi-level categorical variables, only age group and SIMD were suitable among the covariates. We expanded the initial age group variable to include five levels: 18-29, 30-49, 50-65, 66-79 and above 79. We first evaluated a mixed-effect model with random intercepts only, which did not positively impact the performance. We then attempted to add various random slope terms interacting with the intercepts for age and SIMD. A chi-squared test based on the AIC scores of the mixed models suggested that models with a random intercept and random slope term performed significantly better than models with only a random intercept term (p<0·001). The best-identified model was achieved using age group as the random slope and SIMD as the random intercept. The opposite case (age group as the random intercept term and SIMD as the random slope term) yielded a high conditional $R^{2}$ but a low marginal $R^{2}$. On the other hand, the former model achieved a considerably improved explained variance compared to the rest of the models, minimal residual error and moderate reliability within the identified clusters, as expressed by the Intra-cluster Correlation Coefficient (ICC).

**Table S3. Results from the regression model sensitivity analysis using an Ordinary Least Squares Regression on different dataset variations and a Linear Mixed Effects Regression incorporating random intercepts and slopes into the existing models.**

| Model variation | | R^2^ (marginal) | R^2^ (conditional) | AIC | BIC | RMSE | ICC |
| --- | --- | --- | --- | --- | --- | --- | --- |
| Ordinary Least Squares Regression | **Baseline** | 0.037 | 0.037 | 31,943 | 32,105 | 0.963 | / |
|  | **Balanced for COVID-19 status** | 0.114 | 0.114 | 62,338 | 62,513 | 1.01 | / |
|  | **Balanced, removed outliers** | 0.146 | 0.146 | 54,169 | 54,342 | 0.941 | / |
|  | **Balanced, removed outliers, stepwise feature selection** | 0.146 | 0.146 | 54,164 | 54,323 | 0.941 | / |
| Linear Mixed Effects Regression* | **I – Age group** | 0.142 | 0.167 | 54,275 | 54,441 | 0.941 | 0.029 |
|  | **I - SIMD** | 0.156 | 0.144 | 54,275 | 54,441 | 0.941 | 0.013 |
|  | **I – Age group, S – COVID-19 status** | 0.287 | 0.728 | 54,166 | 54,348 | 0.938 | 0.577 |
|  | **I – Age group, S – SIMD** | 0.113 | 0.846 | 54,102 | 54,379 | 0.935 | 0.826 |
|  | **I – Age group, S – Sex** | 0.125 | 0.739 | 54,206 | 54,387 | 0.939 | 0.703 |
|  | **I – SIMD, S – COVID-19 status** | 0.162 | 0.146 | 54,279 | 54,461 | 0.941 | 0.019 |
|  | **I – SIMD, S – Age group** | **0.363** | **0.822** | **54,116** | **54,392** | **0.935** | **0.721** |
|  | **I – SIMD, S – Sex** | 0.302 | 0.709 | 54,222 | 54,403 | 0.94 | 0.583 |

Model terms: I – random intercept, S – random slope

Metrics: AIC – Akaike Information Criterion, BIC – Bayesian Information Criterion, RMSE – Root Mean Squared Error, ICC – Intraclass Correlation Coefficient;

* The linear mixed effects regression model was fit on the balanced COVID-19 cohort, trimmed for outliers using the covariates selected from the stepwise feature selection.

#### 7. **Random effect parameters from optimal fit**

The overall performance measures relative to the random effects of the optimal LMER model fit are highlighted in Table S4. The model sample size included the reduced set of observations after outlier removal. There were five cluster combinations in the model relative to the SIMD quintiles. The total residual error *σ^2^* indicated the mean random effect variance across all combinations. The between-subject (random intercept) variance (τ_00_) indicated the variance across SIMD group interactions. In contrast, the random slope variance (τ_11_) specified the detected variance in interactions between SIMD and each respective age group, where the youngest group was treated as the reference. The amount of explained variance between the random effect terms was strongest within the 66-79 age group (τ_11_=2·7) compared to the other interactions. This was due to the significantly higher density within this age group, leading to the strongest area of adjustment. The incorporated mixed effects structure with random intercepts and slopes explained the overall proportion of the variance (ICC=0·72) with moderate reliability. Additionally, the total residual error (σ^2^=0·88) represented a reasonable measure of fit. Regarding potential signs of heteroscedasticity, the null hypothesis was not rejected (p=0·052). There were also no identified signs of overfitting relating to model singularity, as the test for boundary fits was negative.^12^

**Table S4. Random effect estimates from the optimal Linear Mixed Effects Regression model using SIMD as the random intercept and age group as the random slope terms.**

| Random effects evaluated for optimal model fit | |
| --- | --- |
| *σ^2^* | 0.88 |
| *τ_00_(SIMD)* | 0.73 |
| *τ_11_(SIMD, Age group [30-49])* | 0.15 |
| *τ_11_(SIMD, Age group [50-65])* | 0.43 |
| *τ_11_(SIMD, Age group [66-79])* | 2.7 |
| *τ_11_(SIMD, Age group [Above 79])* | 0.36 |
| *ICC* | 0.72 |
| *N_SIMD_* | 5 |
| *Observations* | 19,922 |
| *Marginal R^2^ / Conditional R^2^* | 0.363 / 0.822 |

Random effect terms: σ^2^ – residual model variance (error); τ_00_ – between-subject (random intercept) variance; τ_11_ – random slope variance; ICC – intra-class correlation coefficient
